# Transcranial random noise stimulation for the acute treatment of depression: a randomized controlled trial

**DOI:** 10.1101/19004218

**Authors:** Stevan Nikolin, Angelo Alonzo, Donel Martin, Veronica Gálvez, Sara Buten, Rohan Taylor, James Goldstein, Cristal Oxley, Dusan Hadzi-Pavlovic, Colleen K. Loo

**Author notes:** **Corresponding Author** Email., Address: Black Dog Institute, Hospital Road, Randwick 2031 NSW, Australia. **Funding** NHMRC project grant APP1051423.

## Abstract

**Background:** Transcranial electrical stimulation has broad potential as a treatment for depression. Transcranial random noise stimulation (tRNS), which delivers randomly fluctuating current intensities, may have greater cortical excitatory effects compared to other forms of transcranial electrical stimulation. We therefore aimed to investigate the antidepressant efficacy of tRNS.

**Methods:** Depressed participants were randomly assigned by computer number generator to receive 20 sessions of either active or sham tRNS over four weeks in a double-blinded, parallel group randomized-controlled trial. tRNS was delivered for 30mins with a direct current offset of 2mA and a random noise range of 2mA. Primary analyses assessed changes in depression severity using the Montgomery-Asperg Depression Rating Scale (MADRS). Neuroplasticity, neuropsychological, and safety outcomes were analysed as secondary measures.

**Results:** 69 participants were randomised, of which three discontinued treatment early leaving 66 (sham n = 34, active n = 32) for per-protocol analysis. Depression severity scores reduced in both groups (MADRS reduction in sham = 7.0 [95%CI 5.0-8.9]; and active = 5.2 [95%CI 3.2-7.3]). However, there were no differences between active and sham groups in the reduction of depressive symptoms, or the number of participants meeting response (sham = 14.7%; active = 3.1%) and remission criteria (sham = 5.9%; active = 0%). Erythema, paraesthesia, fatigue, and dizziness/light-headedness occurred more frequently in the active tRNS group. Neuroplasticity, neuropsychological and acute cognitive effects were comparable between groups.

**Conclusion:** Our results do not support the use of tRNS with the current stimulation parameters as a therapeutic intervention for the treatment of depression.

Clinical trial registration at clinicaltrials.gov/NCT01792414.

**Significance Statement:** This is the first randomized sham-controlled clinical trial of a four-week course of transcranial random noise stimulation (tRNS) for the treatment of depression. tRNS is a relatively novel form of non-invasive electrical stimulation that uses mild, randomly fluctuating currents to constrain homeostatic mechanisms and increase brain excitability. We investigated effects across multiple validated mood outcomes and comprehensively assessed cognitive, neurophysiological, and physical side effects to examine the safety of tRNS. We found no differences between active and sham conditions for all mood outcomes, and are thus unable to lend support for tRNS as an effective treatment for depression. We found tRNS to be well-tolerated with no adverse acute cognitive, neuropsychological or severe phyisical side effects, suggesting a course of 20 repeated sessions can be delivered safely.

## Introduction

Although there are a number of established treatments for depression, a sizeable proportion of patients still fail to adequately respond, with a conservative estimate of approximately one-third of these not reaching remission even after four trials of different antidepressant medication classes (A J Rush et al., 2006b). In addition, many patients fail to complete a course of antidepressants due to side effects (A John Rush et al., 2006a; Trivedi et al., 2006). While electroconvulsive therapy (ECT) remains the most effective treatment, with response rates of up to 70% (Haq et al., 2015), treatment uptake and adherence can be limited by patient concerns over possible cognitive side effects and the need for general anaesthetic. Thus, there has been interest in the development of novel, non-convulsive brain stimulation techniques that are well-tolerated and have a benign side effect profile, such as transcranial electrical stimulation. These neuromodulatory techniques could have the greatest potential for translation into widespread clinical use, being relatively inexpensive, easy to use, portable, and safe (Bikson et al., 2016; Nikolin et al., 2018). Here we report an investigation of the efficacy of one such technique, transcranial random noise stimulation (tRNS), for the treatment of depression.

Transcranial electrical stimulation involves applying a weak electrical current to cerebral tissue via scalp electrodes, resulting in modulation of neuronal membrane potentials and spontaneous firing rates (M. A. Nitsche et al., 2008) that can lead to long term changes in cortical excitability and plasticity (Michael Nitsche and Paulus, 2001; Michael J Player et al., 2014). Applying a direct current between the electrodes, referred to as transcranial direct current stimulation (tDCS), has been demonstrated to have antidepressant effects in clinical trials (Colleen K Loo et al., 2012; Andre R Brunoni et al., 2013a; Andre R Brunoni et al., 2017). Recent meta-analyses of randomised, sham-controlled trials have found tDCS to be more effective than sham stimulation, with significantly higher remission and response rates as well as a greater reduction in depressive symptoms (André R. Brunoni et al., 2016; Mutz et al., 2018). As depression has increasingly been conceptualised as a disorder underpinned by disrupted neuroplasticity (Pittenger and Duman, 2008; Liu et al., 2017), cumulative changes to synaptic functioning may underlie the therapeutic effects observed in clinical trials of tDCS (Szymkowicz et al., 2016). Modifying stimulation parameters to enhance cortical excitability effects may therefore present a pathway to increase treatment efficacy.

tRNS is a more recently developed transcranial electrical stimulation technique that involves randomly fluctuating current intensities over a broad frequency spectrum (between 0.1 to 640 Hz). There is some evidence that tRNS has comparable, if not greater, cortical excitatory effects as compared to tDCS (Moliadze et al., 2014b; Ho et al., 2015; Inukai et al., 2016b). A single session of 10 minutes of 1 milliampere (mA) tRNS to the motor cortex has been found to produce a greater increase in cortical excitability than tDCS, lasting up to 60 minutes beyond the stimulation period (Moliadze et al., 2014a; Inukai et al., 2016a), although tDCS may lead to a longer period of excitation of at least 90 minutes post-stimulation (Moliadze et al., 2014a). tRNS may also be applied with a direct current offset so that the stimulation incorporates neuromodulatory features of tDCS, in addition to limiting homeostatic responses via randomly fluctuating current intensities. Results from Ho et al. (2015) suggest that tRNS with a direct current offset may be more effective in increasing motor cortical excitability than the more common application of tRNS without an offset.

There has been a burgeoning growth in recent years of studies investigating the application of tRNS to enhance sensory processing (Ghin et al., 2018; K. S. Rufener et al., 2018; Contemori et al., 2019), motor performance (Abe et al., 2019; Jooss et al., 2019), and cognition (Snowball et al., 2013; Popescu et al., 2016; Mammarella et al., 2017; Shalev et al., 2018; Tyler et al., 2018) in healthy participants with largely promising results. To date, however, few studies have examined the effectiveness of tRNS for clinical/therapeutic uses, with such studies typically characterised by small sample sizes and/or varying efficacy (Chan et al., 2012; Haesebaert et al., 2014; U. Palm et al., 2016; Hayward et al., 2017; Kreuzer et al., 2017; Salemi et al., 2019). Regarding the effects of tRNS on mood, there is currently only one report involving treatment of major depressive disorder (MDD). Chan et al. (2012) reported a case of a patient diagnosed with MDD who had responded to two trials of tDCS (2 mA, 20 minutes, 15 sessions over 3 weeks) prior to trialling a 4-week course of open-label tRNS (2 mA range with 1 mA direct current offset, 20 sessions lasting 20 minutes each). It was found that by the 15^th^ session, there was a 63% reduction from baseline in the severity of depressive symptoms, compared with a reduction of 31% and 25% at the end of the acute treatment phase in the two prior trials of tDCS. For all three trials, depression scores at baseline were similar but the patient reported faster improvement with tRNS and lesser skin sensations compared to tDCS. Given this encouraging case report finding, and the potential theoretical advantages of tRNS relative to tDCS, further investigation of the antidepressant effects of tRNS is warranted.

The primary aim of this study, therefore, was to conduct the first randomised, sham-controlled trial of tRNS in depression. It was hypothesised that tRNS would have significant antidepressant efficacy compared with a sham control over a 4-week treatment phase. A secondary aim of this study was to examine whether antidepressant effects of tRNS were mediated by restoration of brain neuroplasticity. We hypothesised that antidepressant response to tRNS would be associated with increased brain plasticity, given prior findings of reduced neuroplasticity in depressed individuals compared to healthy matched controls (Michael J Player et al., 2013), and findings suggesting a normalisation of neuroplasticity following antidepressant treatment using tDCS (Michael J Player et al., 2014). Lastly, as this is the first treatment trial of tRNS for depression, a comprehensive neuropsychological test battery was designed specifically to be sensitive to symptom changes, measure any adverse cognitive effects, and to assess any potential acute cognitive enhancing effects.

## Materials and Methods

### Trial design

The main study phase used a double-blinded, parallel, randomized, sham-controlled design. Participants were assigned by a computer-generated random number sequence to one of two groups: active tRNS or sham tRNS. Randomization was stratified according to whether participants were diagnosed with unipolar or bipolar depression. Participants were required to attend a total of 20 tRNS sessions over four weeks conducted on consecutive weekdays during the sham-controlled phase. Participants who missed five or more sessions during the sham-controlled phase were withdrawn from the trial, and were excluded from analyses using a per-protocol approach. All participants were offered an additional 20 sessions of open-label active tRNS over four weeks, also administered every weekday. After treatment in the acute daily treatment phases, participants entered a taper phase during which they received once weekly tRNS treatment for four weeks with the final taper session coinciding with a 1-month follow-up visit. Participants were then followed up at 3, 6 and 9 months. Participants and raters were blinded to tRNS condition and blinding was maintained until the study was completed and the dataset locked.

Mood, neuroplasticity, and neuropsychological function were assessed at the intervals shown in Supplementary Table S1. Adequacy of blinding to treatment was assessed at the end of the sham-controlled phase by asking participants and raters to guess the tRNS condition administered during the first four weeks of treatment. To investigate whether treatment expectations may be a predictor of response, participants completed the Treatment Expectancy Questionnaire (TEQ; see Supplementary Figure S1) at baseline before the first tRNS session.

The study was powered for the primary aim of testing efficacy over the sham-controlled phase. From pilot data, it was assumed that tRNS would be at least as effective as tDCS when tested in a sham-controlled trial, given that sampling criteria were very similar. Means and standard deviations of the active and sham treatment groups from our previous, sham-controlled, 3-week trial of tDCS (Colleen K Loo et al., 2012) were used, with outcomes extrapolated for a 4-week comparison period. This resulted in an effect size of Cohen’s *d* = 0.7. For 80% power and α = 0.05, a sample of 33 subjects per group was required to demonstrate a difference between active and sham treatment.

### Participants

At study entry, participants were at least 18 years old; in a current major depressive episode (as part of a MDD or Bipolar Disorder) of a minimum four weeks duration, defined according to Diagnostic and Statistical Manual of Mental Disorders (fourth edition, text revision; DSM-IV-TR) criteria and established using the Mini International Neuropsychiatric Interview (MINI; Version 5.0.0) (Sheehan et al., 1998) and study clinician assessment; and had a total score of at least 20 on the Montgomery-Asberg Depression Rating Scale (MADRS) (Montgomery and Asberg, 1979). Participants were free of antidepressant medications, or continued on stable doses of antidepressant medications to which they had failed to respond after an adequate course of treatment, with dosage unchanged for at least four weeks prior to study entry. Bipolar participants were required to be on a mood stabilizer medication (e.g. lithium, valproate, or carbamazepine) as prophylaxis against treatment-emergent mania or hypomania for the duration of the study.

Exclusion criteria included: psychotic disorder as per DSM-IV-TR; drug or alcohol abuse or dependence within 12 months of study entry; inadequate response to ECT in the current depressive episode; current benzodiazepine medication; rapid clinical response required (e.g., due to high suicide risk); clinically defined neurological disorder or insult; metal in the cranium; skull defects; skin lesions on the scalp at electrode sites; or pregnancy.

The study was approved by the human research ethics committee of the University of New South Wales and was conducted at the Black Dog Institute in Sydney, Australia. Participants provided written informed consent for this study. Recruitment began in January 2013 and the last follow-up was conducted in 2017. The study was registered with the ClinicalTrials.gov website (Identifier: NCT01792414).

### Transcranial random noise stimulation

A DC-Stimulator Plus device (NeuroConn GmbH, Germany) applied high frequency tRNS (100-640 Hz) via two 7 × 5 cm saline-soaked sponge-covered electrodes held in position by a headband. Active tRNS was administered for 30 mins per session with a range of 2 mA and an offset of 2 mA. The anode was placed over F3 (as per the 10-20 international electroencephalogram system), corresponding to the left dorsolateral prefrontal cortex (LDLPFC), and the cathode over F8. For sham stimulation, the current was ramped up over 10 s, left on for 30 s, then gradually ramped down over 10 s, so that both treatment groups experienced an initial tingling sensation. The tRNS machine was then left on until the end of the session to preserve blinding. This sham procedure resulted in adequate blinding for tDCS in previous trials (e.g., (C. K. Loo et al., 2010; Colleen K Loo et al., 2012)), and was therefore expected to be sufficient for tRNS, which produces milder skin sensations compared to tDCS (Ambrus et al., 2010). Participants were comfortably seated at rest and did not engage in any particular tasks during stimulation.

### Clinical outcome measures

The primary outcome measure for comparing active and sham tRNS was the MADRS, which was administered by trained raters with established inter-rater reliability (intraclass correlation coefficient > 0.7). Secondary measures were the Clinician Global Impression – Improvement (CGI-I) (Guy, 1976), Beck Depression Inventory (BDI-II) (Beck et al., 1996) and Quality of Life Enjoyment and Satisfaction Questionnaire – Short Form (Q-LES-SF) (Endicott et al., 1993) scales.

### Neuroplasticity outcome measures

As an optional study offered to participants in the main tRNS trial, a paired associative stimulation (PAS) paradigm previously described in M. J. Player et al. (2012) was used to assess the effects of tRNS on neuroplasticity. The PAS testing was conducted at baseline before the first tRNS session, and again after completion of the sham-controlled and open-label phases. Briefly, the PAS paradigm involves measuring motor evoked potentials (MEPs) following single-pulse transcranial magnetic stimulation (TMS) via electromyography (EMG) before and after applying a stimulation protocol (i.e., PAS) to the motor cortex to assess changes in motor cortical excitability (see the Supplementary Material for a detailed description of PAS methodology).

### Neuropsychological outcome measures

The following neuropsychological battery was administered to comprehensively assess cognitive function: California Verbal Learning Test-II (CVLT-II) (Delis et al., 2000) – verbal learning and memory; Ruff 2 & 7 (Ruff and Allen, 1996) – attention processes; Wechsler Adult Intelligence Scale-IV edition (WAIS-IV) Digit Span subtest (Wechsler, 2008) – simple auditory attention and working memory; Symbol Digit Modality Test (SDMT) (Smith, 1991) – psychomotor processing speed; Delis-Kaplan Executive Function System (D-KEFS) Verbal Fluency test (Delis et al., 2001) – phonemic fluency, semantic fluency, cognitive flexibility; and Cognitive Failures Questionnaire (Broadbent et al., 1982) – subjective cognitive functioning. Alternative versions of the CVLT-II, D-KEFS Verbal Fluency, and SDMT were used to minimize practice effects. In addition, computer administered cognitive tests were used to assess safety and acute effects. A simple reaction time test, in which participants were instructed to press a space bar as soon as they saw a cross appear in the middle of a computer screen, was administered immediately before and after the first tRNS session. An Emotion Recognition Task (Montagne et al., 2007), which assessed recognition of six basic facial emotions, was also administered after the first tRNS session.

### Physical adverse events

As an additional safety outcome measure, physical adverse events were assessed each session using a tRNS Side Effects Questionnaire (Supplementary Figure S2), adapted from Andre Russowsky Brunoni et al. (2011), which collected information regarding the type of adverse event, its severity, and its causality.

### Statistical Analyses

All statistical analyses were conducted using SPSS software (IBM SPSS Statistics 25 for Windows; SPSS Inc.). Outcome measures were analysed for change over the sham-controlled phase using a mixed-effects repeated measures (MERM) model with a restricted number of covariates. Time was entered as a repeated measures factor with an unstructured covariance matrix, tRNS Condition (active or sham) was a between-subjects factor, and subjects were included as a random effect. For mood and quality of life outcomes, covariates were selected based on prior reports of their significant effect on antidepressant response to transcranial electrical stimulation; these included treatment resistance (Bikson et al., 2016) (assessed by the Maudsley Staging Method (Fekadu et al., 2009)), and presence of concurrent antidepressant medications e.g. selective-serotonin reuptake inhibitors, serotonin– norepinephrine reuptake inhibitors (A. R. Brunoni et al., 2013b; Andre R Brunoni et al., 2013a). A MERM model was similarly used for neuropsychological outcomes, with MADRS mood scores at the respective time points included as a covariate. Acute cognitive effects following the first session were examined using a two-way repeated measure analysis of variance (RMANOVA), with factors of tRNS Condition and Time (pre and post session 1). For the Emotion Recognition Task, a multivariate ANOVA (MANOVA) was conducted with the between-subject factor of tRNS Condition.

Additional MERM analyses were conducted for the primary outcome measure (MADRS). Baseline scores on the TEQ were added as a covariate to the MERM analysis to test whether treatment expectations modified mood outcomes. To assess whether medication use affected outcomes, each medication class (antidepressants, benzodiazepines, antipsychotics, lithium and anticonvulsants) was entered as the only covariate in separate MERM analyses.

The number of responders (defined as a reduction in MADRS total score of ≥ 50% from baseline) and remitters (defined as a final MADRS total score < 10) at the end of the sham-controlled phase were compared between active and sham tRNS groups using a Fisher’s exact test.

The association between participant or rater guesses (active or sham) and the participant’s assigned tRNS condition (active or sham) was tested using a Pearson Chi-square test with Yates’ continuity correction. Cohen’s kappa statistic was used to assess agreement between participant and rater guesses.

Statistical tests were two-tailed and significance was set at *p* < 0.05.

## Results

A total of 69 participants met inclusion criteria and were randomized to receive either active or sham tRNS during the sham-controlled phase (see the CONSORT flow diagram, Supplementary Figure S3). A total of 66 participants (sham: 34, active: 32) completed the sham-controlled phase and were analysed using a per-protocol approach. Table 1 shows demographic and clinical characteristics for active and sham tRNS groups at baseline.

**Table 1.** Comparison of demographic and clinical characteristics at baseline.

|  | Sham | Active |
| --- | --- | --- |
| n | 34 | 32 |
| *Medications (yes/no)* |  |  |
| Any concurrent medication | 27/7 | 22/10 |
| Antidepressant | 23/11 | 20/12 |
| Lithium | 1/33 | 2/30 |
| Benzodiazepine* | 2/32 | 2/30 |
| Antipsychotic | 6/28 | 4/28 |
| Anticonvulsant | 5/29 | 1/31 |
| *Clinical and demographic variables (m, SD)* |  |  |
| Gender (m/f) | 19/15 | 17/15 |
| Melancholic (yes/no) | 19/9 | 12/12 |
| MDD/BP1/BP2 | 30/3/1 | 30/0/2 |
| Age (years) | 48.8 (12.3) | 47.5 (12.0) |
| Age at onset (years) | 27.0 (9.6) | 27.5 (9.6) |
| Duration of current episode (months) | 24.4 (32.2) | 37.2 (48.0) |
| Duration of previous episodes (months) | 65.9 (65.8) | 88.5 (93.6) |
| Antidepressants failed current episode | 2.1 (1.7) | 2.6 (2.9) |
| Total lifetime failed antidepressants | 4.0 (3.0) | 5.0 (5.1) |
| Maudsley Staging score | 7.3 (2.1) | 6.5 (1.9) |
| Baseline MADRS score | 29.5 (4.6) | 30.1 (5.0) |
| Baseline BDI-II score | 33.8 (9.1) | 34.1 (9.4) |
| Baseline CGI-S score | 4.4 (0.6) | 4.4 (0.6) |
| Baseline Q-LES-SF score | 32.8 (10.5) | 34.9 (11.2) |
| Baseline TEQ score | 24.5 (7.6) | 24.1 (7.1) |

\*Participants were required to cease benzodiazepine medication use prior to commencing the trial.
MDD: Major depressive disorder. BP1/BP2: Bipolar I and bipolar II disorder. MADRS: Montgomery-Asberg Depression Rating Scale. Q-LES-SF: Quality of Life Enjoyment and Satisfaction Questionnaire – Short Form. BDI-II: Beck Depression Inventory. CGI-S: Clinician Global Impression Severity scale, is a 7-point clinician-rated scale ranked from 1-normal to 7-extremely ill. TEQ: Treatment Expectancy Questionnaire.

### Clinical outcome measures

Table 2 shows the results for all MERM analyses of mood and quality of life outcome measures during the sham-controlled phase using per-protocol and intention-to-treat approaches in accordance with CONSORT guidelines for parallel group randomised control trials (Schulz et al., 2010). MERM analysis of MADRS scores showed a significant effect of Time (*p* < 0.001). There was, however, no effect of tRNS Condition (*p* = 0.630), and no significant Time × Condition interaction (*p* = 0.445; see Figure 1). Repeating the analysis while incorporating scores from the Treatment Expectations Questionnaire as an additional covariate did not modify outcomes. Concurrent medications did not significantly affect outcomes when entered as covariates in separate MERM analyses (see Supplementary Table S2). Supplementary Table S3 shows results from MERM analyses of all outcome measures acquired during the acute daily treatment phases combined (i.e. both sham-controlled and open-label phases). Supplementary Figure S4 shows graphs of mood and quality of life measures for all time-points up to the 9-month follow-up assessment.

**Table 2.** Mood and quality of life outcome measures. Estimated marginal means and results from MERM analyses during the sham-controlled phase. MERM analyses were performed including the following covariates: Maudsley staging method total score as a measure of treatment resistance, and antidepressant use. Statistically significant p-values (<0.05) are highlighted in bold.

| *Mood Assessments - mean (n, SEM)* | Baseline |  | Week 2 |  | Week 4 |  | Condition |  | Time |  | Time × Condition |  |
| --- | --- | --- | --- | --- | --- | --- | --- | --- | --- | --- | --- | --- |
|  | Sham | Active | Sham | Active | Sham | Active | F | p | F | p | F | p |
| Per-protocol analysis |  |  |  |  |  |  |  |  |  |  |  |  |
| MADRS | 29.1 (33, 0.8) | 28.9 (32, 0.8) | 24.4 (33, 1.0) | 24.8 (32, 1.1) | 22.1 (34, 1.2) | 23.7 (32, 1.3) | 0.24 | 0.630 | 41.73 | **<0.001** | 0.82 | 0.445 |
| BDI-II | 32.6 (31, 1.7) | 33.2 (28, 1.8) | 25.2 (34, 1.9) | 27.2 (32, 2.0) | 22.1 (32, 1.9) | 24.9 (31, 2.0) | 0.72 | 0.399 | 24.56 | **<0.001** | 0.46 | 0.762 |
| CGI-I | - | - | 3.5 (33, 0.1) | 3.6 (32, 0.1) | 3.1 (33, 0.1) | 3.4 (32, 0.1) | 2.24 | 0.140 | 12.69 | **<0.001** | 2.10 | 0.153 |
| Q-LES-SF | 32.4 (34, 1.9) | 36.0 (30, 2.1) | - | - | 43.5 (32, 2.8) | 43.9 (31, 2.9) | 0.45 | 0.503 | 31.67 | **<0.001** | 0.84 | 0.362 |
| Intention-to-treat analysis |  |  |  |  |  |  |  |  |  |  |  |  |
| MADRS | 29.1 (34, 0.8) | 29.0 (34, 0.8) | 24.4 (33, 1.0) | 24.8 (34, 1.1) | 22.1 (34, 1.2) | 23.7 (32, 1.3) | 0.29 | 0.593 | 43.09 | **<0.001** | 0.77 | 0.467 |
| BDI-II | 32.4 (32, 1.6) | 33.4 (30, 1.7) | 24.9 (34, 1.9) | 27.1 (34, 2.0) | 21.9 (32, 1.9) | 25.1 (31, 2.0) | 1.07 | 0.306 | 25.07 | **<0.001** | 0.58 | 0.678 |
| CGI-I | - | - | 3.5 (33, 0.1) | 3.6 (34, 0.1) | 3.1 (33, 0.1) | 3.4 (32, 0.1) | 2.60 | 0.112 | 13.07 | **<0.001** | 1.98 | 0.165 |
| Q-LES-SF | 32.5 (35, 1.8) | 35.8 (32, 2.0) | - | - | 43.6 (32, 2.8) | 43.8 (31, 2.9) | 0.37 | 0.545 | 32.00 | **<0.001** | 0.80 | 0.376 |

MADRS: Montgomery-Asberg Depression Rating Scale. BDI-II: Beck Depression Inventory. CGI-I: Clinician Global Impression Improvement scale, is a 7-point clinician-rated scale ranked from 1-very much improved to 7-very much worse. Q-LES-SF: Quality of Life Enjoyment and Satisfaction Questionnaire – Short Form. m: Mean. SEM: Standard error of measure.

**Figure 1.**
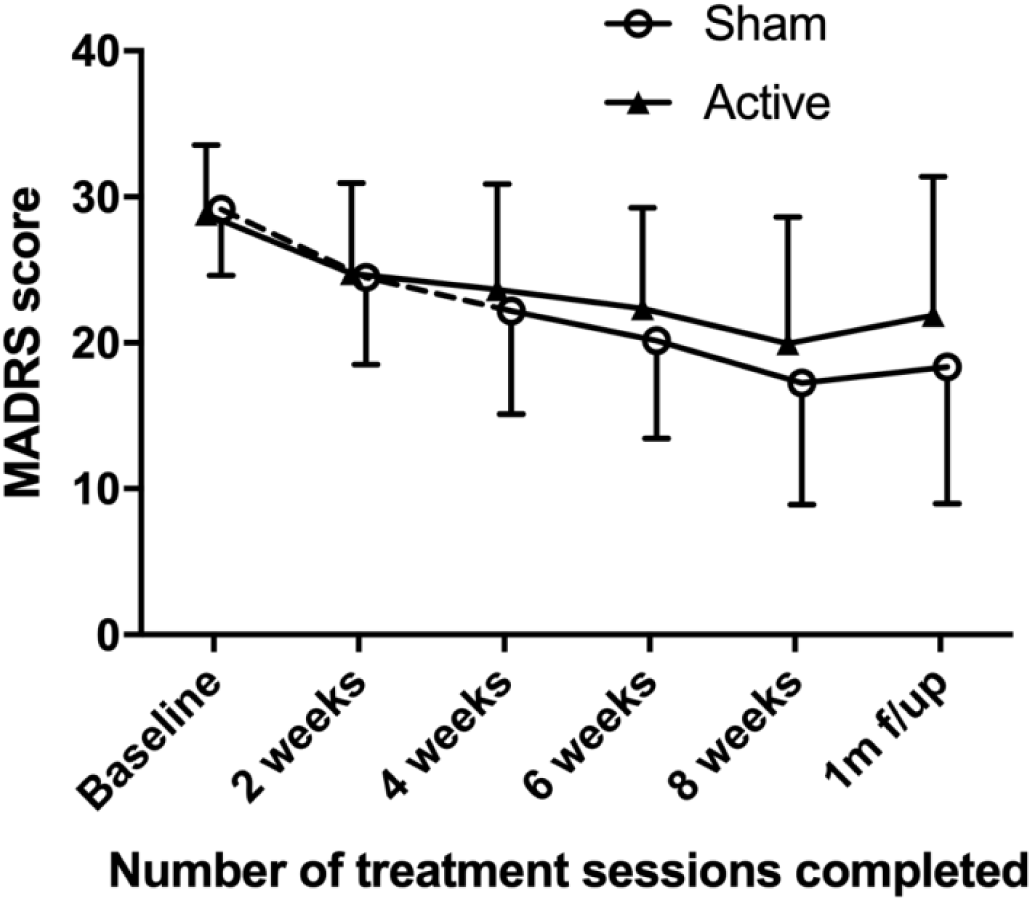
Mood scores. Graph showing MADRS scores (estimated marginal means ± SD) across rating time points, including the sham-controlled phase (from baseline to 4-weeks), open-label phase (from 4 – 8 weeks), and 1-month follow-up assessment following the final taper session. Dotted lines indicate sham tRNS sessions delivered during the sham-controlled phase.

One participant in the active tRNS group (3.1%), and five participants receiving sham tRNS (14.7%), were considered treatment responders after completion of the sham-controlled phase (i.e. following 20 sessions of tRNS). Only two participants met the remission criterion, both in the sham tRNS condition (5.9%). Fisher’s exact tests revealed no statistically significant differences between active and sham tRNS conditions for response (*p* = 0.198) and remission (*p* = 0.493) rates.

### Neuroplasticity outcome measures

A total of 44 participants (sham: 25, active: 19) completed the optional PAS study to assess changes in motor cortical excitability. A MERM analysis found no significant effects of Time (*p* = 0.209) or Condition (*p* = 0.780), and no significant Time × Condition interaction (*p* = 0.570; see Table 3, and Supplementary Figure S5). Furthermore, change in MADRS scores did not correlate with changes in MEP amplitudes from baseline to the end of the sham-controlled phase (r = -0.02, *p* = 0.905; Supplementary Figure S6). Results from intention-to-treat analyses are also reported in Supplementary Table S4.

**Table 3.** Neuroplasticity and neuropsychological outcome measures. Estimated marginal means and results from MERM analyses during the sham-controlled phase. The neuroplasticity MERM analysis were performed including the following covariates: Maudsley staging parameters total score as a measure of treatment resistance, and antidepressant use. Neuropsychological MERM analyses were performed using MADRS scores as a covariate. Statistically significant p-values (<0.05) are highlighted in bold.

|  | Baseline |  | Week 4 |  | Condition |  | Time |  | Time × Condition |  |
| --- | --- | --- | --- | --- | --- | --- | --- | --- | --- | --- |
|  | Sham | Active | Sham | Active | F | p | F | p | F | p |
| *Neuroplasticity Assessment (m, SEM)* |  |  |  |  |  |  |  |  |  |  |
| PAS: MEP amplitude | 1.24 (0.10) | 1.21 (0.12) | 1.31 (0.09) | 1.40 (0.10) | 0.08 | 0.780 | 1.63 | 0.209 | 0.33 | 0.570 |
| *Neuropsychological Assessments (m, SEM)* |  |  |  |  |  |  |  |  |  |  |
| CVLT-II: Trial 1-5 total recall t-score | 45.7 (2.8) | 48.7 (2.2) | 46.3 (2.3) | 49.7 (2.2) | 2.12 | 0.148 | 0.12 | 0.734 | 0.01 | 0.913 |
| CVLT-II: Long delay free recall z-score | -0.41 (0.21) | 0.03 (0.22) | -0.64 (0.22) | -0.38 (0.21) | 2.69 | 0.104 | 1.85 | 0.176 | 0.20 | 0.653 |
| D-KEFS: Letter fluency scaled score | 11.2 (0.7) | 11.5 (0.7) | 10.9 (0.7) | 11.9 (0.7) | 0.97 | 0.327 | 0.01 | 0.945 | 0.32 | 0.575 |
| D-KEFS: Category fluency scaled score | 10.3 (0.8) | 10.9 (0.8) | 9.8 (0.8) | 11.8 (0.8) | 2.84 | 0.094 | 0.05 | 0.828 | 0.80 | 0.372 |
| D-KEFS: Category switching total scaled score | 10.0 (0.6) | 11.1 (0.6) | 9.6 (0.6) | 10.6 (0.6) | 3.36 | 0.069 | 0.50 | 0.480 | 0.02 | 0.888 |
| Ruff 2 & 7: Total speed t-score | 47.4 (1.9) | 49.9 (1.9) | 51.3 (1.9) | 53.6 (1.8) | 1.66 | 0.200 | 3.45 | 0.066 | 0.01 | 0.946 |
| Ruff 2 & 7: Total accuracy t-score | 49.6 (2.3) | 46.6 (2.4) | 50.4 (2.4) | 48.7 (2.3) | 1.02 | 0.316 | 0.34 | 0.561 | 0.08 | 0.779 |
| SDMT: Total correct z-score | -0.31 (0.18) | 0.25 (0.18) | -0.02 (0.19) | 0.41 (0.18) | 7.78 | **0.006** | 1.32 | 0.254 | 0.15 | 0.703 |
| WAIS-IV Digit Span: Total correct scaled score | 9.7 (0.5) | 11.5 (0.6) | 10.5 (0.6) | 12.6 (0.5) | 13.00 | **<0.001** | 2.57 | 0.112 | 0.12 | 0.732 |
| CFQ: Total score | 50.0 (2.2) | 46.8 (2.3) | 49.9 (2.3) | 50.0 (2.2) | 0.52 | 0.474 | 0.40 | 0.528 | 0.57 | 0.451 |

PAS: Paired associative stimulation. MEP: motor evoked potential. CVLT: California Verbal Learning Test. WAIS: Wechsler Adult Intelligence Scale. SDMT: Symbol Digit Modalities Test. D-KEFS: Delis-Kaplan Executive Function System. CFQ: Cognitive Failures Questionnaire. m: Mean. SEM: Standard error of measure.

### Neuropsychological outcomes

Neuropsychological outcomes during the sham-controlled phase are shown in Table 3. There were no significant main effects of Time, and no significant Time × Condition interaction effects for all neuropsychological measures. Results from intention-to-treat analyses are also reported in Supplementary Table S4.

### Acute cognitive effects

For reaction time, the main effects of Time (*p* = 0.404), Condition (*p* = 0.992), and the Time × Condition interaction effect (*p* = 0.949) were not statistically significant. Further, for the Emotion Recognition Task the main effect of tRNS Condition was not statistically significant (*p* = 0.347).

### Physical adverse events

Adverse events occurring during the sham-controlled and open-label phases of the trial are presented in Table 4. Side-effects were transient and mild-to-moderate in severity. Pearson Chi-square tests revealed significantly more instances of erythema (skin redness; *p* < 0.001), paraesthesia (tingling, burning, and itching sensations; *p* < 0.001), fatigue (*p* = 0.006), and dizziness/light-headedness (*p* = 0.002) following active tRNS as compared to sham tRNS sessions.

**Table 4.**
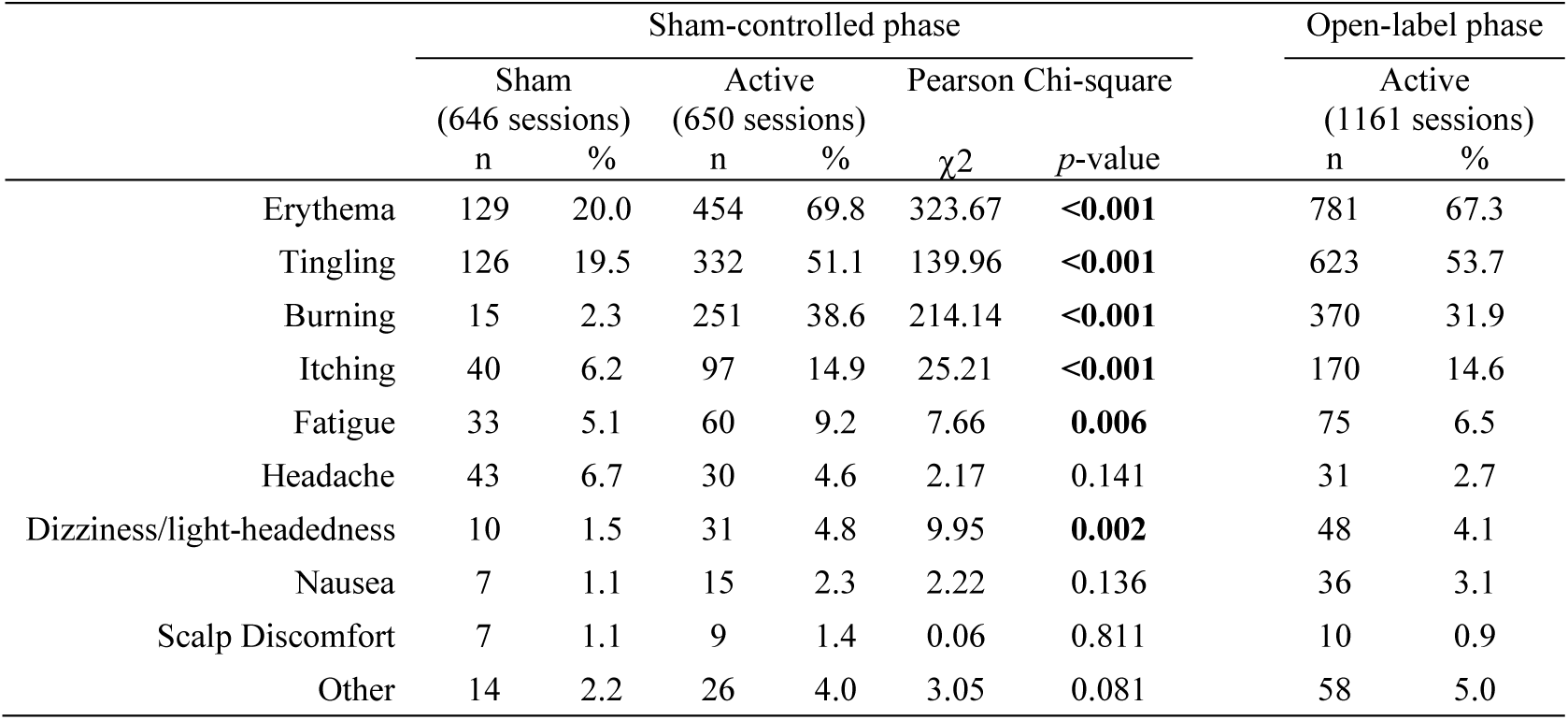
Physical adverse event frequency. Adverse events are sorted according to overall likelihood of occurrence, with events most likely to occur listed first. Differences in frequency of adverse event occurrence during the sham-controlled phase was tested using Pearson Chi-square tests with Yates’ continuity correction. Statistically significant p-values (<0.05) are highlighted in bold.

### Blinding integrity

Participants were asked to guess their treatment condition at the end of the double-blinded sham-controlled phase. 75% of participants in the sham condition correctly guessed they received sham tRNS, and 55% of participants in the active condition correctly guessed they had received active tRNS. A Pearson Chi-square test of participant guesses was significant (χ^2^ = 4.68; *p* = 0.031), suggesting that participants were not adequately blinded to their treatment condition. To determine whether participant guesses of treatment condition may have influenced mood outcomes, we performed a post-hoc simple linear regression analysis; percent change in MADRS score over the 4-week sham-controlled phase was selected as the dependent variable, and participant guess (active, sham) as the independent variable. This analysis was not statistically significant (R^2^ = 0.023, F = 1.43, p = 0.237).

Blinded study raters were similarly asked to guess participants’ treatment condition at the end of the sham-controlled phase. Raters correctly guessed that participants had received sham tRNS 56% of the time, and active tRNS 31% of the time. A Pearson Chi-square test found this difference not to be statistically significant (χ^2^ = 0.492; p = 0.483). Furthermore, Cohen’s kappa showed no agreement between participant and rater guesses (κ = 0.134; p = 0.329).

## Discussion

Here we report the results of the first randomized control trial to examine the efficacy of tRNS for the treatment of depression. Although there was a significant reduction of depressive symptoms over the duration of the study period, there was no difference in the rate of improvement between sham and active tRNS conditions. Further, there was no significant effect of tRNS on neuroplasticity measures in the subset of participants that completed the PAS paradigm, suggesting that stimulation did not increase global cortical excitability. tRNS was found to be safe with no adverse acute cognitive, neuropsychological or severe phyisical side effects. However, tRNS resulted in a higher incidence rate of skin redness (erythema) and paraesthesia (tingling, itching, and burning sensations) in the active condition, as well as fatigue and dizziness/light-headedness, which occurred in fewer than 10% of sessions. Nevertheless, the stimulation protocol was well-tolerated with only one participant dropping-out due to adverse effects, which were not conclusively associated with tRNS.

The results of this study do not support the use of tRNS with the current stimulation parameters as a therapeutic intervention for the treatment of depression. Despite encouraging initial evidence of significant reductions in depression scores in patients with fibromyalgia (Curatolo et al., 2017), and a case report of improvement in MDD (Chan et al., 2012), mood and quality of life outcomes in the active tRNS group were no different from the placebo-controlled response in the sham group at all time-points. The size of reductions in depressive symptoms observed in both conditions of the present study is broadly similar to the sham condition of previous trials of tDCS for depression (Blumberger et al., 2012; Colleen K Loo et al., 2012;U Palm et al., 2012; Andre R Brunoni et al., 2013a; Bennabi et al., 2015; Sampaio-Junior et al., 2018). Of these previous tDCS trials, our study design was most similar to a recent international multisite investigation conducted by our group, which observed improvements in depression scores of 27.8% and 22.3% in sham and active tDCS conditions, respectively, as compared to 24.1% and 18.0% in sham and active tRNS conditions (Colleen K Loo et al., 2018). We recruited participants using analogous inclusion/exclusion criteria, participant demographic and clinical characteristics, and adopted comparable stimulation parameters, at least in terms of treatment duration (30 minutes), session number and frequency (20 daily weekday sessions), and direct current intensity (2 mA direct current offset in the present study *vs* a marginally stronger current intensity of 2.5 mA in the tDCS study). Interestingly, the present tRNS trial and Colleen K Loo et al. (2018) have used the highest total number of sessions and strongest stimulation parameters, including current intensity, compared to other investigations of the antidepressant effects of transcranial electrical stimulation (Blumberger et al., 2012; Colleen K Loo et al., 2012; U Palm et al., 2012; Andre R Brunoni et al., 2013a; Bennabi et al., 2015), with both studies reporting no advantage of active stimulation over sham. Overall, studies show tDCS has antidepressant efficacy (Mutz et al., 2018), though one study suggested this may be less than escitalopram (Andre R Brunoni et al., 2017).

Another possible explanation for null findings in the present study is that the sample was too severely ill to respond to tRNS treatment. On average, participants had failed 4-5 antidepressants and depression scores just below the cut-off for severe depression (MADRS > 34; see Table 1). Research suggests that participants with treatment resistant depression do not respond as well transcranial electrical stimulation, such as tDCS (Blumberger et al., 2012; U Palm et al., 2012; Bennabi et al., 2015).

The synaptic plasticity hypothesis of depression purports that MDD is characterized by a partial reduction in long-term potentiation-like processes (Michael J Player et al., 2013; Kuhn et al., 2016), suggesting that impaired synaptic plasticity, particularly in the prefrontal cortex, is a key feature of the pathophysiology of depression (Goto et al., 2010; Duman et al., 2016; Noda et al., 2018). This hypothesis is supported by evidence that the therapeutic efficacy of antidepressants is, at least partly, due to their capacity to increase neural plasticity (Santarelli et al., 2003; Castrén and Hen, 2013). Similarly, prior work from our group has demonstrated that a course of tDCS increases neuroplasticity and mood outcomes in depression. However, a correlation between these measures could not be confirmed due to the limited sample size (n = 18) (Michael J Player et al., 2014). Despite a larger sample size from which to detect small effects of the intervention, we did not see an increase in neuroplasticity levels following tRNS. Previous research has shown that tRNS can induce acute neuroplastic after-effects, measured by investigating changes to motor cortex excitability using TMS motor evoked potentials (Terney et al., 2008; Chaieb et al., 2015; Ho et al., 2015). However, there is no evidence to date to suggest cumulative changes in neuroplasticity following a course of multiple repeated sessions of tRNS, as assessed in this study. An important caveat to our results is that only a subset of participants (44/69) completed the PAS study. It is theoretically possible that the subset of participants who underwent the PAS protocol had different clinical, physiological, or behavioural characteristics (e.g. greater levels of motivation) and may thus not be representative of the larger sample.

The exact purported mechanisms of action for prolonged cortical excitation following tRNS are unclear, but may include either 1) temporal summation of neural activity when random noise stimuli and ongoing endogenous neuronal activity occur in close succession (Fertonani et al., 2011), and/or 2) enhancement of neuronal signalling via the principle of stochastic resonance (van der Groen and Wenderoth, 2017). The latter refers to signals that are too weak to exceed a threshold being amplified by adding a random noise stumulus, improving the signal-to-noise ratio and the synchronization and coherence of neuronal networks (Moss et al., 2004; Pavan et al., 2019). Though more research is required to determine which of these mechanisms dominates, the action of tRNS appears to rely heavily on detection and propagation of weak ongoing endogenous neuronal signals. The notion of stochastic resonance has been demonstrated in several tRNS experiments aimed at enhancing sensitivity to sensory inputs, including visual, auditory, and tactile stimuli (van der Groen and Wenderoth, 2017). Interestingly, investigations of auditory (Katharina S Rufener et al., 2017) and visual (Van der Groen et al., 2018) perceptual thresholds have shown that tRNS has its largest effect on near-threshold stimuli, whereas stimuli clearly above and below threshold were unaffected. It may be for this reason that the only positive randomized control trials using tRNS in clinical populations have stimulated the sensory and motor cortices, specifically in the treatment of tinnitus via the auditory cortex (Vanneste et al., 2013), or stimulation of the motor cortex for chronic pain in fibromyalgia (Curatolo et al., 2017). tRNS may restore the dysfunctional activity in these cortical structures by normalizing their capacity to filter weak signals amidst background neural noise. In complex disorders such as depression, however, it is unclear what the depressed ‘signal’ might be. Studies seeking to use tRNS for other complex disorders by stimulating prefrontal cortical regions have also reported negative findings, i.e. for the treatment of multiple sclerosis (U. Palm et al., 2016), and vegetative state (Mancuso et al., 2017). Similarly to depression, these illnesses do not consist of a well-defined neural signal whose signalling properties can be augmented by tRNS to revert pathophysiological dysfunctions of brain activity.

Although the present study reports null findings for the use of tRNS in depression, it provides valuable information regarding the safety and tolerability of multiple repeated sessions of tRNS. To the best of our knowledge, the previous longest delivery of tRNS was 15 sessions (Chan et al., 2012), whereas participants in the current study experienced up to 40 sessions over eight weeks if allocated to the active tRNS condition. Adverse events of erythema and paraesthesia were reported more frequently in the active tRNS condition as compared to sham. These results are comparable to those observed in the tDCS literature, in which meta-analyses similarly suggest a greater frequency of erythema and paraesthesia (Moffa et al., 2017; Nikolin et al., 2018). Of interest, fatigue and dizziness/light-headedness were also noted more frequently during active tRNS sessions. These side effects are not a common adverse effect of tDCS and might be unique to the tRNS stimulation parameters used in the current study, for example, due to the current intensity ranging as high as 3 mA (2 mA direct offset with ±1 mA amplitude fluctuation). Importantly, fatigue and dizziness/light-headedness occurred rarely, in only 9.2% of sessions for fatigue, and 4.8% for light-headedness. Additionally, these adverse events were transient, resolved on their own shortly after cessation of stimulation, and were not reported to be severe in intensity.

A limitation of the present study is that blinding was not preserved, possibly due to the increased incidence of adverse events during active tRNS compared to sham. One would expect inadequate blinding to reduce placebo effects for participants in the sham condition, and potentially enhance them for participants receiving active tRNS, thereby increasing the likelihood of observing a difference between groups. This was not the case in the present study. Indeed, scores for mood outcomes were quantitatively (but not significantly) better in the sham condition compared to active tRNS, suggesting that inadequate blinding did not bias results in favour of the active treatment. Future studies may consider alternative methods to adequately blind participants, including the use of a topical salve beneath the site of stimulation to reduce paraesthetic effects and erythema (McFadden et al., 2011; Guarienti et al., 2015), or comparisons against an active control condition (Fonteneau et al., 2019).

A major strength of the present study is the research design, which included double blinding of participants and raters, examination of mood outcomes in addition to neuroplasticity changes, comprehensive assessment of adverse events using neuropsychological and physical measures, reporting of long-term follow-up outcomes up to 9-months following completion of the open-label phase, and rigorous statistical analysis methodology informed by CONSORT guidelines.

## Conclusion

This study represents the first randomised control trial for the use of tRNS to treat depression. Our findings do not lend support for the use of tRNS as a therapeutic intervention for depression. Antidepressant response was similar between active and sham tRNS conditions. tRNS did not increase motor cortical excitability, a measure of neuroplasticity associated with antidepressant response in other successful therapeutic clinical trials of depression (Santarelli et al., 2003; Castrén and Hen, 2013). The profile of adverse events for tRNS was similar to that of tDCS, with a significantly greater likelihood of erythema and paraesthesia in the active tRNS condition, in addition to a higher incidence rate of fatigue and dizziness/light-headedness. Participant blinding was not preserved and may be related to the increased frequency of side effects in the active condition. Overall, the treatment was well-tolerated by participants.

## Data Availability

Participant consent was not obtained to make data publically available. However, syntax and output from statistical analyses can be found at: https://github.com/snikolin/tRNSdepressionRCT

## Supplementary Materials

### Paired associative stimulation methods

Participants sat in a comfortable, relaxed position with both hands on a pillow. The optimal site for eliciting motor evoked potentials (MEPs) in the right first dorsal interosseous (FDI) muscle was first established and marked on the scalp. Each participant’s resting motor threshold (RMT) – defined as the minimum stimulus intensity to evoke MEPs of ⩾50 μV in the relaxed FDI in 5 out of 10 consecutive trials of single-pulse TMS – was then measured. TMS intensity for evoking test MEPs was adjusted individually for each participant to elicit an average 1 mV response. TMS intensity for testing MEPs was limited to a maximum of 130% of RMT even if the actual intensity needed to elicit a 1 mV response was greater.

For the stimulation protocol, single-pulse TMS (130% of RMT) to the FDI representation on the left motor cortex was combined with electrical stimuli (200 μs duration, 300% perceptual threshold, DS7 stimulator; Digitimer Co. Ltd., Hertfordshire, UK) to the right ulnar nerve proximal to the wrist. Two hundred pairs of stimuli (TMS and ulnar nerve) were given at 0.25 Hz over ∼13 minutes. In each pair, ulnar nerve stimulation preceded TMS by 25 ms. Electrical stimuli were delivered occasionally to the right index finger during PAS. Participants counted the stimuli and reported the number at the end of the session to ensure sensory attention to the hand (Stefan et al., 2004). The MEPs recorded before and after PAS to assess cortical excitability consisted of testing blocks of 20 MEPs at a rate of 0.1 Hz. Two blocks of MEPs were recorded at baseline. A block of MEPs was recorded immediately after PAS and then every 10 mins for one hour. During all stimulation, EMG from the FDI was monitored to ensure muscle relaxation. MEPs were discarded if EMG showed substantial muscle activity (i.e. peak-to-peak amplitude ≥ 50 μV) in the time window prior to the TMS stimuli. Additionally, block-averaged MEP amplitudes were considered outliers if they were more than 4 standard deviations from the mean for active and sham tRNS conditions combined.

EMG activity was recorded through Ag/AgCl surface electrodes over the right FDI. EMG was amplified, bandpass filtered (16–1000 Hz), and digitized (2000 Hz) (Cambridge Electronics Design, Cambridge, UK). TMS (Magstim 200 stimulator, Magstim Co., Whitland, UK) was applied using a 70-mm figure-of eight coil oriented with the handle posterolateral, 45° to the parasagittal plane.

**Supplementary Table S1.** Schedule of assessments.

|  | Baseline | Sham-controlled phase |  |  |  | Open-label phase |  |  |  | Taper phase |  |  |  |
| --- | --- | --- | --- | --- | --- | --- | --- | --- | --- | --- | --- | --- | --- |
| Week | Wk 0 | Wk 1 | Wk 2 | Wk 3 | Wk 4 | Wk 1 | Wk 2 | Wk 3 | Wk 4 | Wk 1 | Wk 2 | Wk 3 | Wk 4 |
| Session | Pre 1 | 1-5 | 6-10 | 11-15 | 16-20 | 21-25 | 26-30 | 31-35 | 36-40 | 41 | 42 | 43 | 44 |
| *Mood Assessments* |  |  |  |  |  |  |  |  |  |  |  |  |  |
| MADRS | X |  | X |  | X |  | X |  | X |  |  |  | X |
| BDI-II | X | X | X | X | X | X | X | X | X | X | X | X | X |
| CGI-S | X |  |  |  |  |  |  |  |  |  |  |  |  |
| CGI-I |  |  | X |  | X |  | X |  | X |  |  |  | X |
| Q-LES-SF | X |  |  |  | X |  |  |  | X |  |  |  | X |
| *Neurophysiological Assessments* |  |  |  |  |  |  |  |  |  |  |  |  |  |
| PAS | X |  |  |  | X |  |  |  | X |  |  |  |  |
| *Neuropsychological Assessments* |  |  |  |  |  |  |  |  |  |  |  |  |  |
| CVLT-II | X |  |  |  | X |  |  |  | X |  |  |  |  |
| Ruff 2 & 7 | X |  |  |  | X |  |  |  | X |  |  |  |  |
| WAIS-IV Digit Span | X |  |  |  | X |  |  |  | X |  |  |  |  |
| SDMT | X |  |  |  | X |  |  |  | X |  |  |  |  |
| D-KEFS Verbal Fluency | X |  |  |  | X |  |  |  | X |  |  |  |  |
| CFQ | X |  |  |  | X |  |  |  | X |  |  |  |  |

Wk: Week. MADRS: Montgomery-Asberg Depression Rating Scale. BDI-II: Beck Depression Inventory. CGI-S: Clinician Global Impression Severity scale, is a 7-point clinician-rated scale ranked from 1-normal to 7-extremely ill. CGI-I: Clinician Global Impression Improvement scale, is a 7-point clinician-rated scale ranked from 1-very much improved to 7-very much worse. Q-LES-SF: Quality of Life Enjoyment and Satisfaction Questionnaire – Short Form. PAS: Paired associative stimulation. CVLT: California Verbal Learning Test. WAIS: Wechsler Adult Intelligence Scale. SDMT: Symbol Digit Modalities Test. D-KEFS: Delis-Kaplan Executive Function System. CFQ: Cognitive Failures Questionnaire

**Supplementary Table S2.** MADRS mixed-effects repeated measures analyses using other covariates. Results from MERM analyses during the sham-controlled phase to examine the impact of medications and treatment expectations as covariates on mood outcomes (i.e. MADRS scores).

|  | Covariate |  | Time |  | Condition |  | Time x Condition |  |
| --- | --- | --- | --- | --- | --- | --- | --- | --- |
|  | F | p | F | p | F | p | F | p |
| Baseline TEQ | 0.28 | 0.599 | 40.93 | <0.001 | 0.99 | 0.325 | 1.32 | 0.276 |
| Antidepressants | 1.14 | 0.289 | 44.28 | <0.001 | 2.03 | 0.160 | 1.11 | 0.335 |
| Lithium | 3.73 | 0.058 | 44.20 | <0.001 | 2.24 | 0.140 | 1.14 | 0.327 |
| Benzodiazepine* | 2.48 | 0.120 | 44.24 | <0.001 | 1.88 | 0.175 | 1.12 | 0.332 |
| Antipsychotics | 1.78 | 0.188 | 44.22 | <0.001 | 2.15 | 0.147 | 1.12 | 0.332 |
| Anticonvulsants | 0.34 | 0.561 | 44.26 | <0.001 | 2.10 | 0.152 | 1.12 | 0.332 |

\*Participants were required to cease benzodiazepine medication use prior to commencing the trial.
TEQ: Treatment Expectations Questionnaire

**Supplementary Table S3.** Mood, quality of life, neuroplasticity and neuropsychological outcome measures. Results from MERM analyses during the acute daily treatment phases combined (i.e. tRNS received during the sham-controlled and open-label phases).

|  | Condition |  | Time |  | Time × Condition |  |
| --- | --- | --- | --- | --- | --- | --- |
|  | F | p | F | p | F | p |
| *Mood Assessments (m, SEM)* |  |  |  |  |  |  |
| MADRS | 1.09 | 0.300 | 29.39 | **<0.001** | 0.75 | 0.560 |
| BDI-II | 2.38 | 0.128 | 17.60 | **<0.001** | 1.00 | 0.447 |
| CGI-I | 6.02 | **0.017** | 15.84 | **<0.001** | 1.72 | 0.173 |
| Q-LES-SF | 0.00 | 0.967 | 30.00 | **<0.001** | 1.39 | 0.257 |
| *Neuroplasticity Assessments (m, SEM)* |  |  |  |  |  |  |
| PAS: MEP amplitude | 0.40 | 0.530 | 2.11 | 0.135 | 0.72 | 0.193 |
| *Neuropsychological Assessments (m, SEM)* |  |  |  |  |  |  |
| CVLT-II: Trial 1-5 total recall t-score | 4.47 | **0.036** | 3.12 | **0.046** | 0.17 | 0.844 |
| CVLT-II: Long delay free recall z-score | 5.40 | **0.021** | 3.03 | 0.051 | 0.23 | 0.799 |
| D-KEFS: Letter fluency scaled score | 2.70 | 0.102 | 0.14 | 0.867 | 0.38 | 0.683 |
| D-KEFS: Category fluency scaled score | 1.97 | 0.165 | 0.74 | 0.477 | 1.69 | 0.190 |
| D-KEFS: Category switching total scaled score | 2.94 | 0.088 | 1.07 | 0.346 | 0.24 | 0.786 |
| Ruff 2 & 7: Total speed t-score | 2.94 | 0.088 | 3.50 | **0.032** | 0.06 | 0.946 |
| Ruff 2 & 7: Total accuracy t-score | 0.80 | 0.372 | 0.73 | 0.482 | 0.22 | 0.805 |
| SDMT: Total correct z-score | 3.58 | 0.063 | 2.80 | 0.065 | 1.11 | 0.335 |
| WAIS-IV Digit Span: Total correct scaled score | 17.17 | **<0.001** | 2.05 | 0.132 | 0.06 | 0.945 |
| CFQ: Total score | 0.20 | 0.659 | 0.12 | 0.891 | 0.45 | 0.639 |

MADRS: Montgomery-Asberg Depression Rating Scale. BDI-II: Beck Depression Inventory. CGI-I: Clinician Global Impression Improvement scale, is a 7-point clinician-rated scale ranked from 1-very much improved to 7-very much worse. Q-LES-SF: Quality of Life Enjoyment and Satisfaction Questionnaire – Short Form. PAS: Paired associative stimulation. MEP: motor evoked potential. CVLT: California Verbal Learning Test. WAIS: Wechsler Adult Intelligence Scale. SDMT: Symbol Digit Modalities Test. D-KEFS: Delis-Kaplan Executive Function System. CFQ: Cognitive Failures Questionnaire. m: Mean. SEM: Standard error of measures

**Supplementary Table S4.**
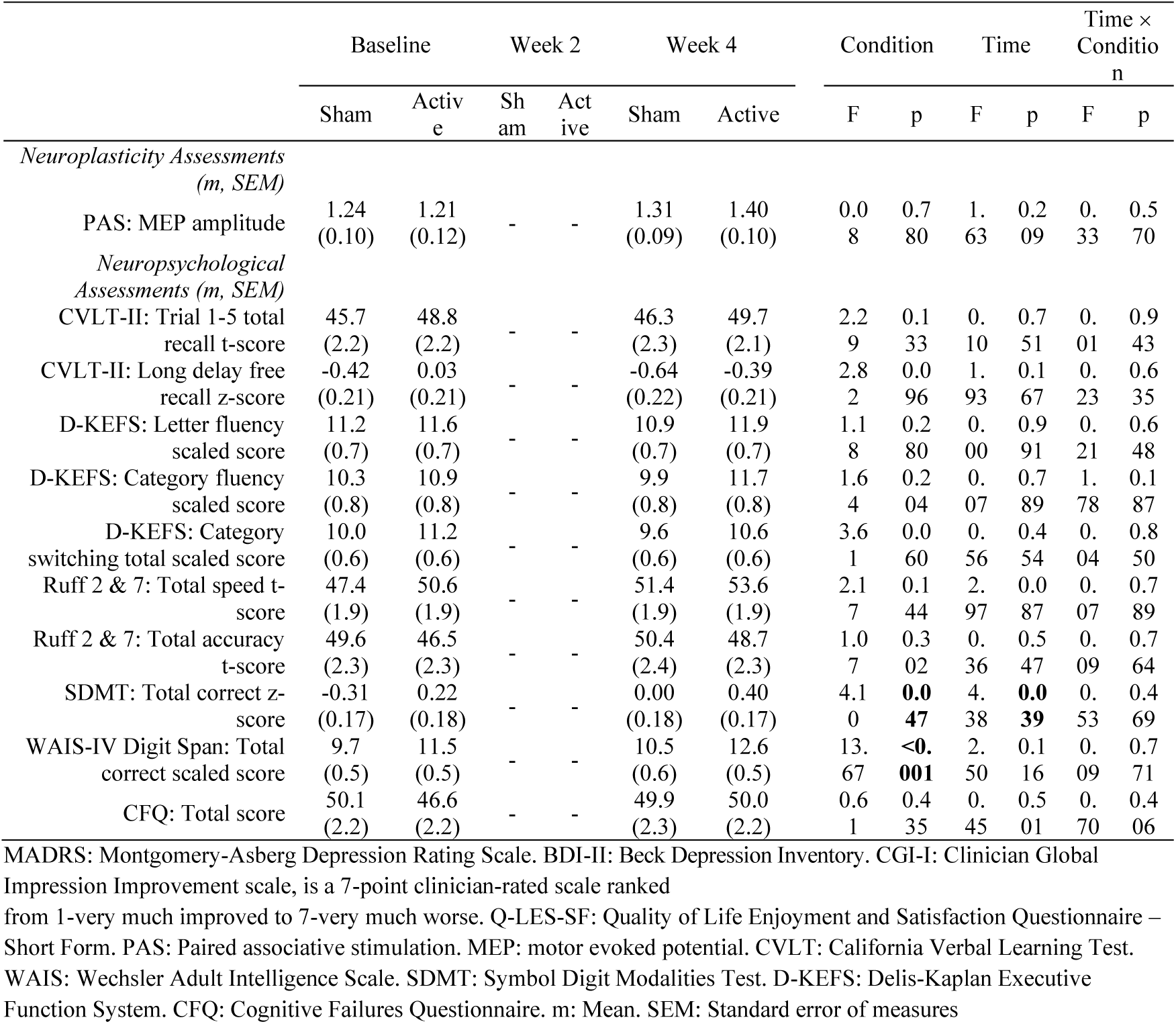
Intention-to-treat analyses. Estimated marginal means and results from MERM analyses during the sham-controlled phase, including the following covariates: Maudsley staging parameters total score as a measure of treatment resistance, and antidepressant use. Neuroplasticity and neuropsychological outcome measures are reported using an intention-to-treat analysis approach.

**Supplementary Figure S1.**
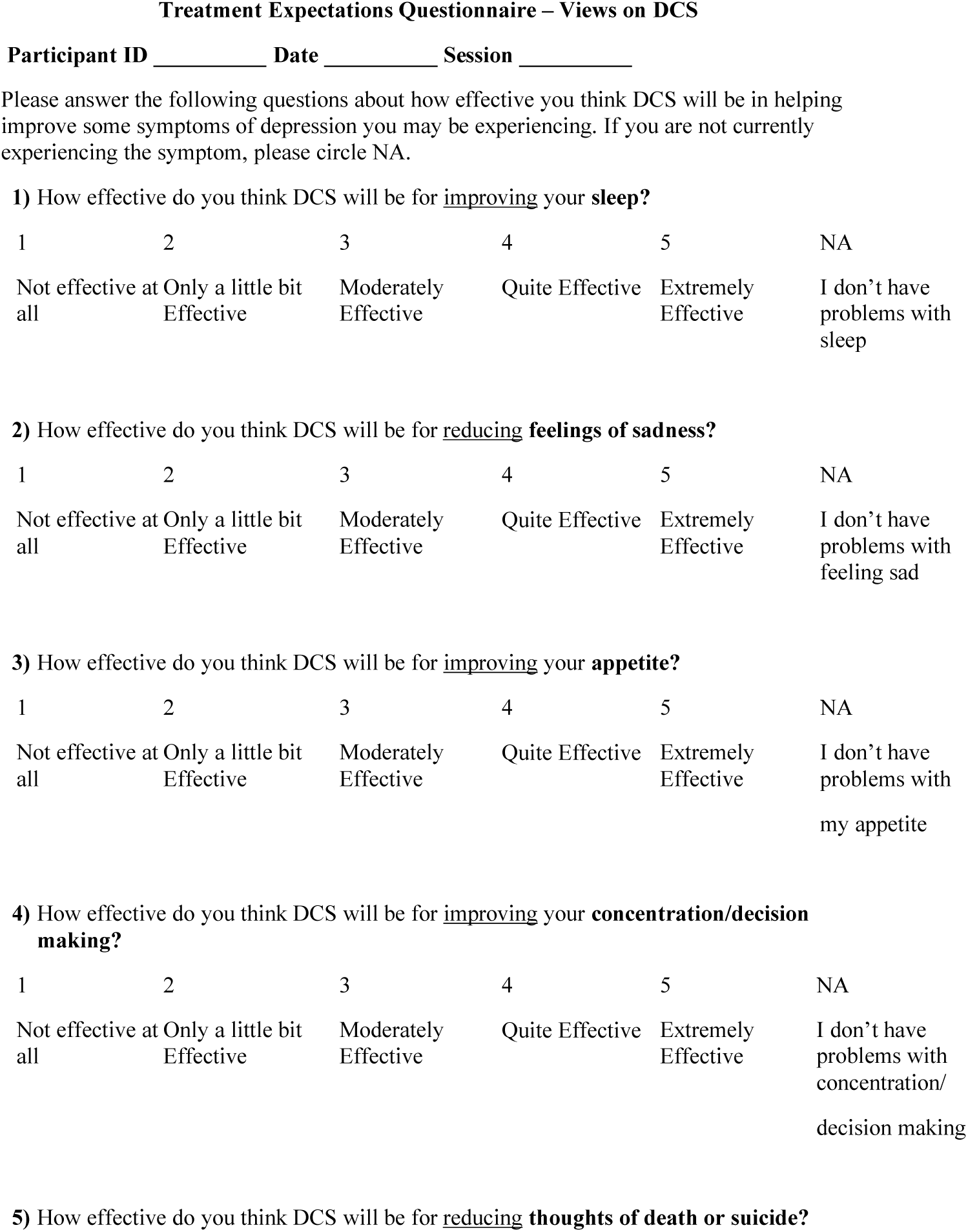
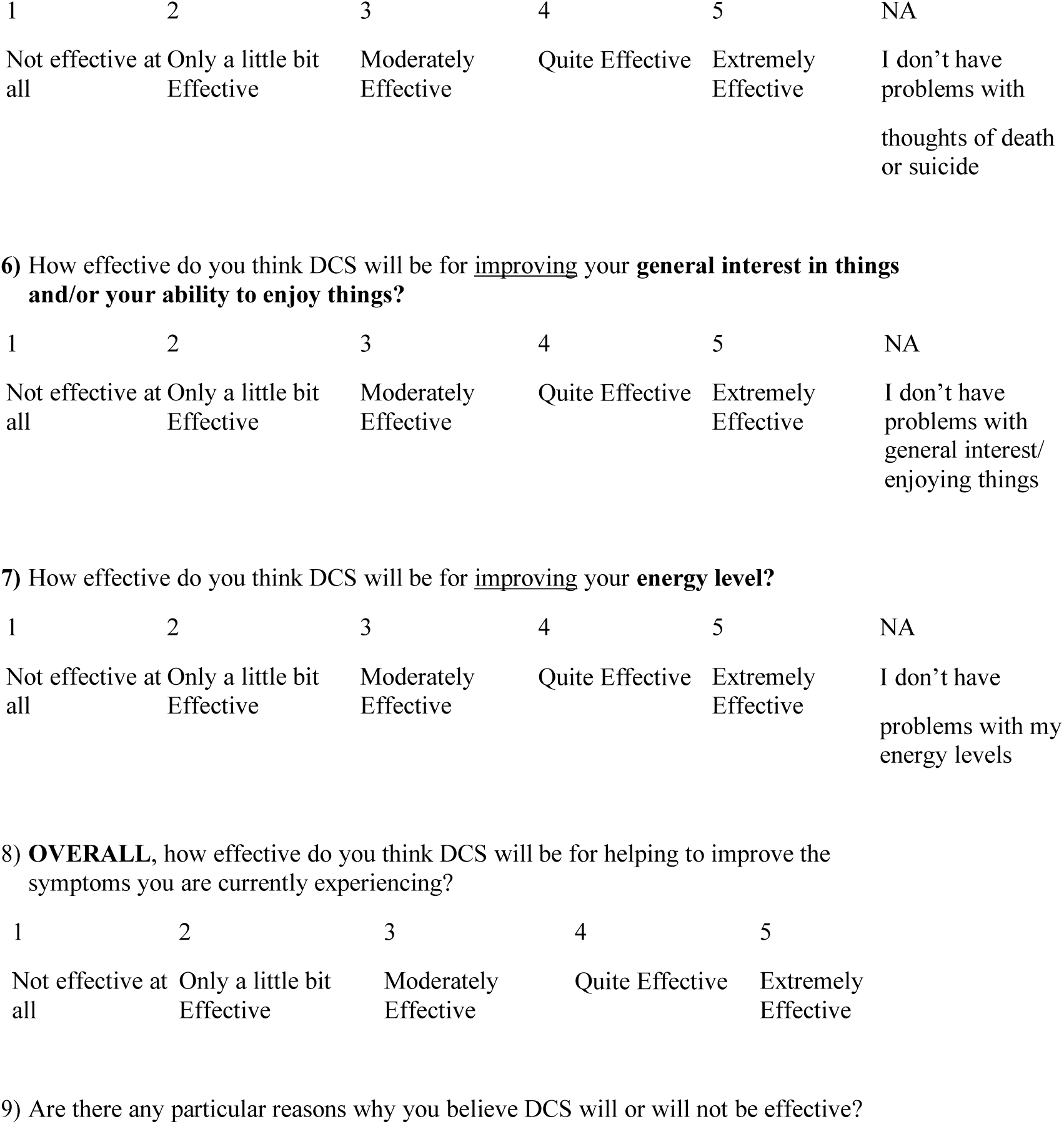
Treatment Expectations Questionnaire. END OF QUESTIONNAIRE

**Supplementary Figure S2.**
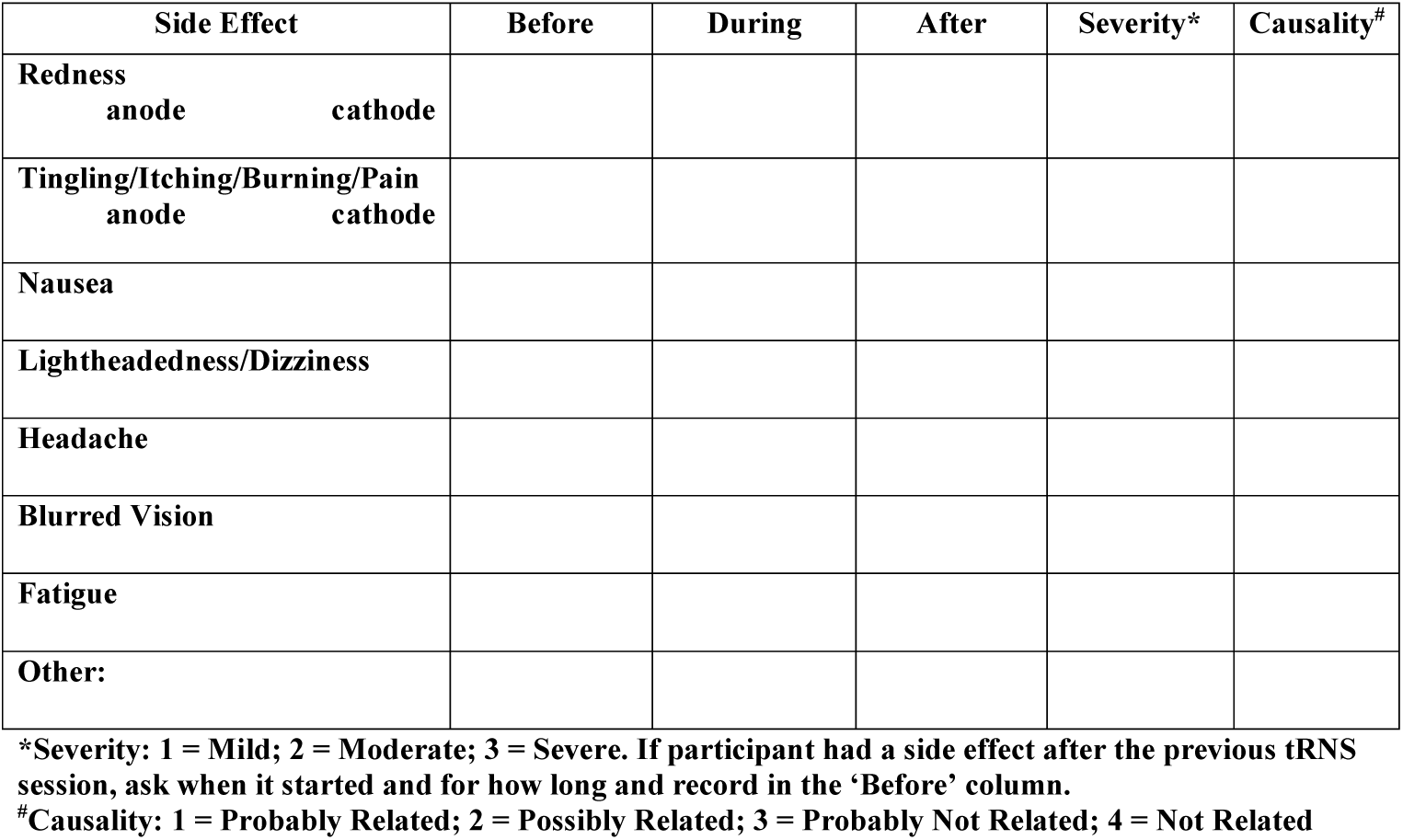
tRNS Side Effects Questionnaire.

**Supplementary Figure S3.**
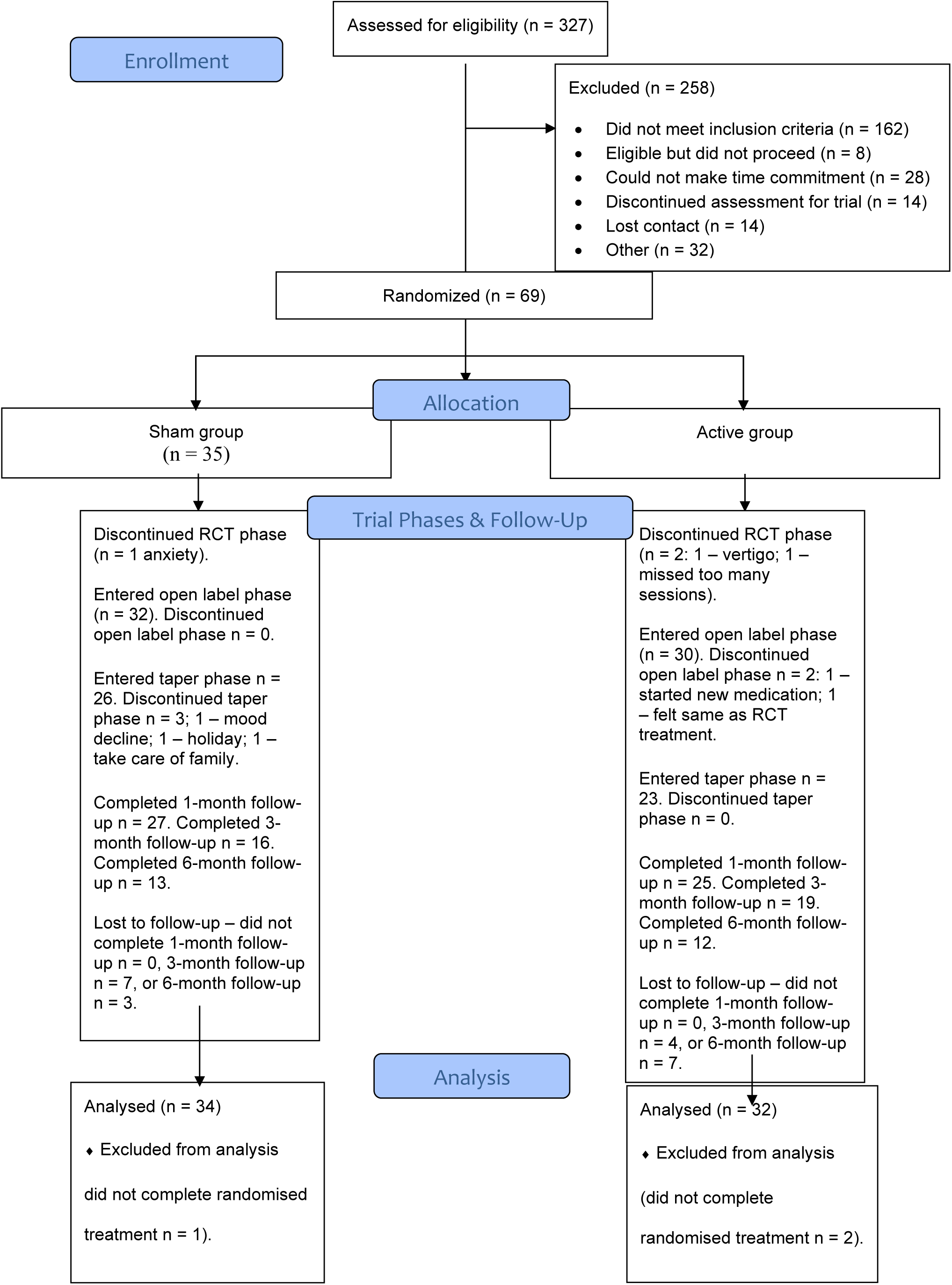
CONSORT flow diagram.

**Supplementary Figure S4.**
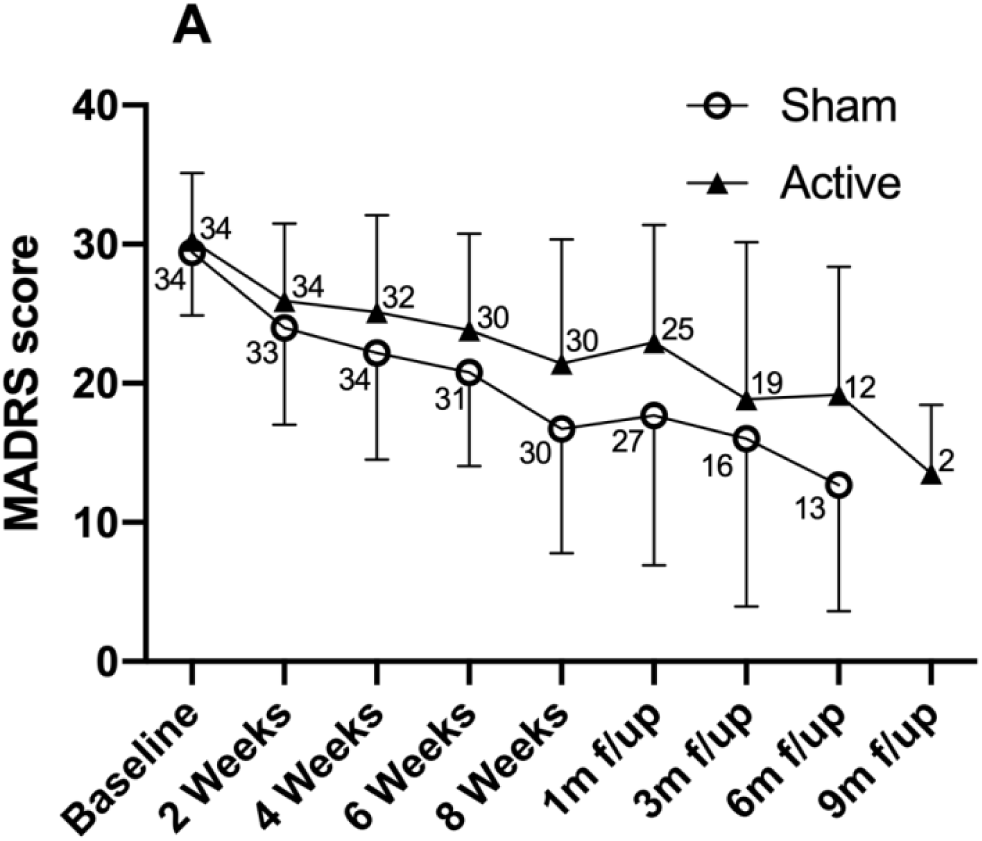
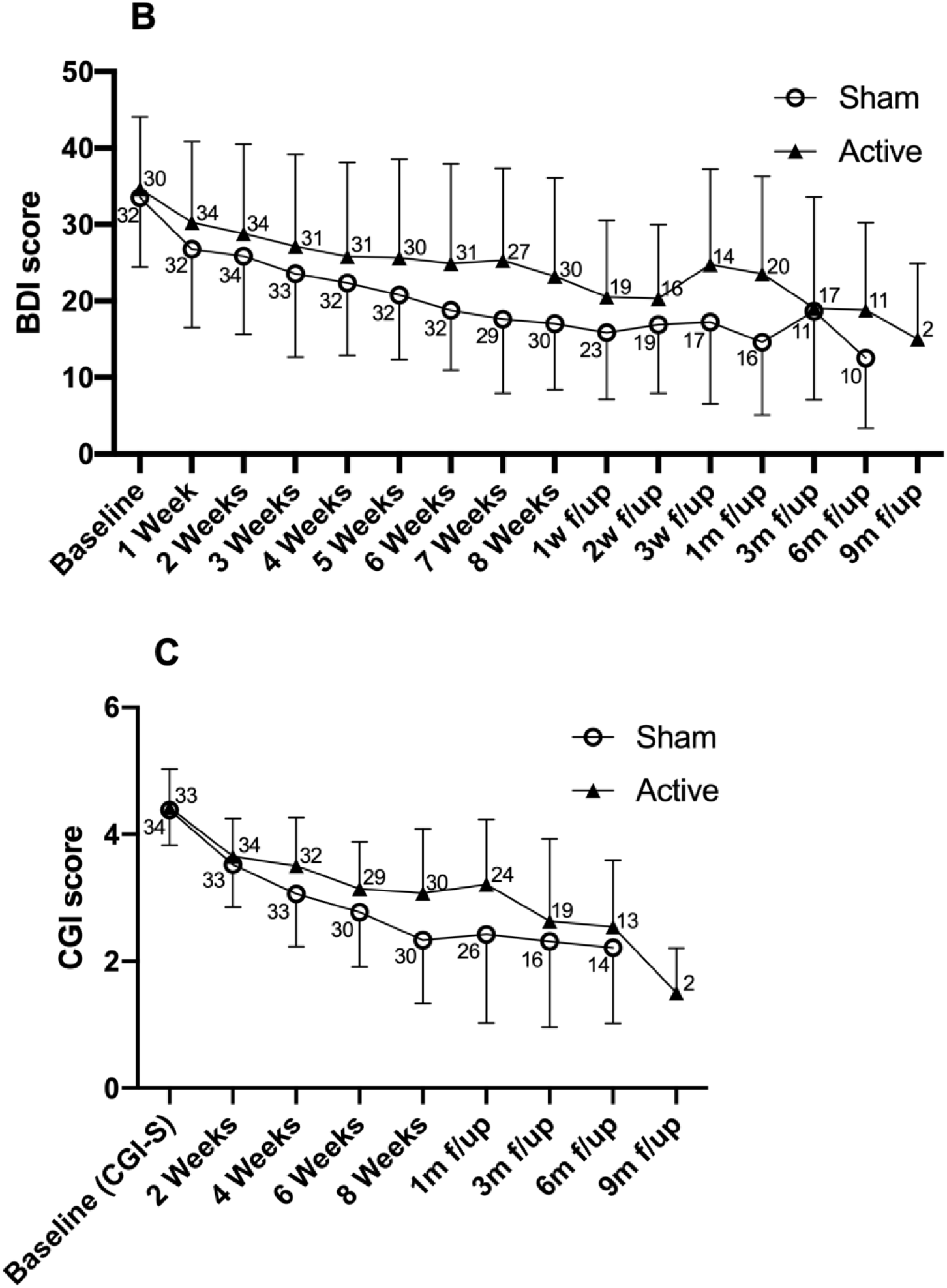
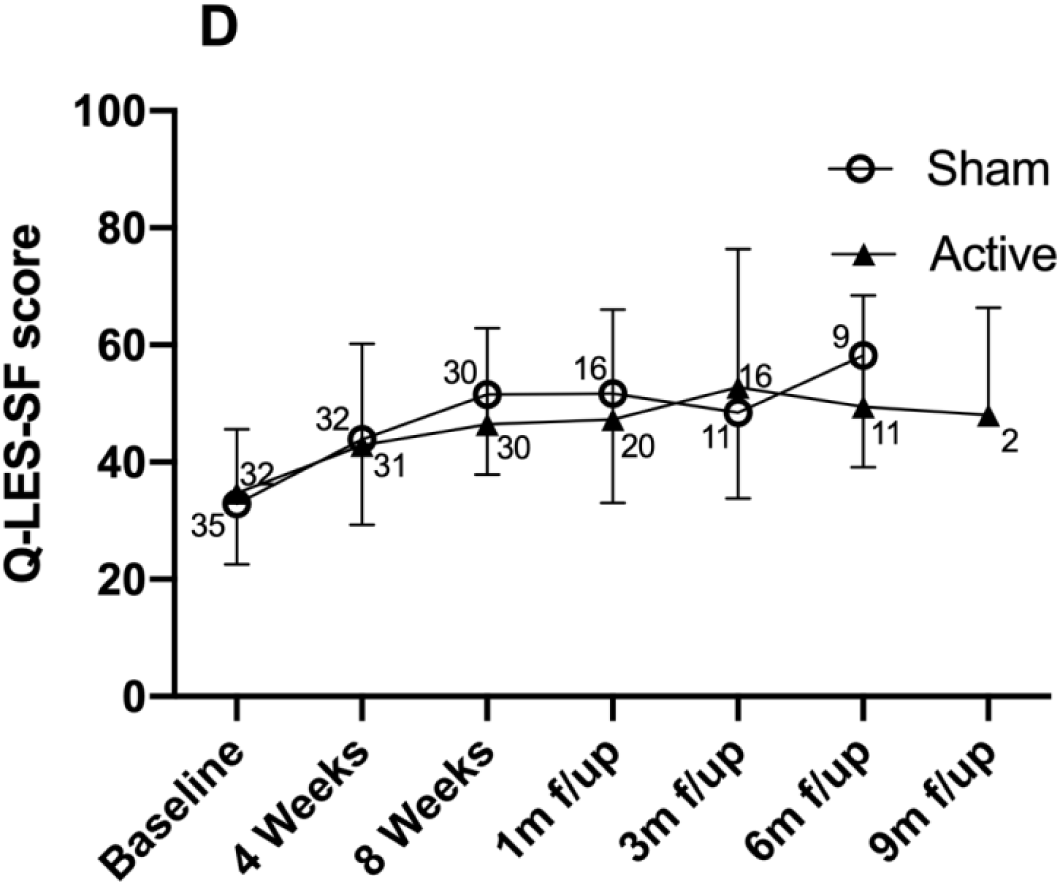
Follow-up mood outcome measures. Graphs showing mood and quality of life scores (unadjusted means ± SD) from baseline up to the 9-month follow-up assessment following completion of the open-label phase. Sample sizes used for the calculation of summary statistics are presented for each time-point. **A)** Montgomery-Asberg Depression Rating Scale (MADRS) scores. **B)** Beck Depression Inventory (BDI-II) scores. **C)** Clinician Global Impression Severity (CGI-S) at baseline, and CGI-Improvement (CGI-I) scores; the CGI-S is a 7-point clinician-rated scale ranked from 1-normal (not at all ill) to 7-among most extremely ill patients. The CGI-I is a 7-point clinician-rated scale ranked from 1-very much improved to 7-very much worse. **D)** Quality of Life Enjoyment and Satisfaction Questionnaire – Short Form scores (Q-LES-SF).

**Supplementary Figure S5.**
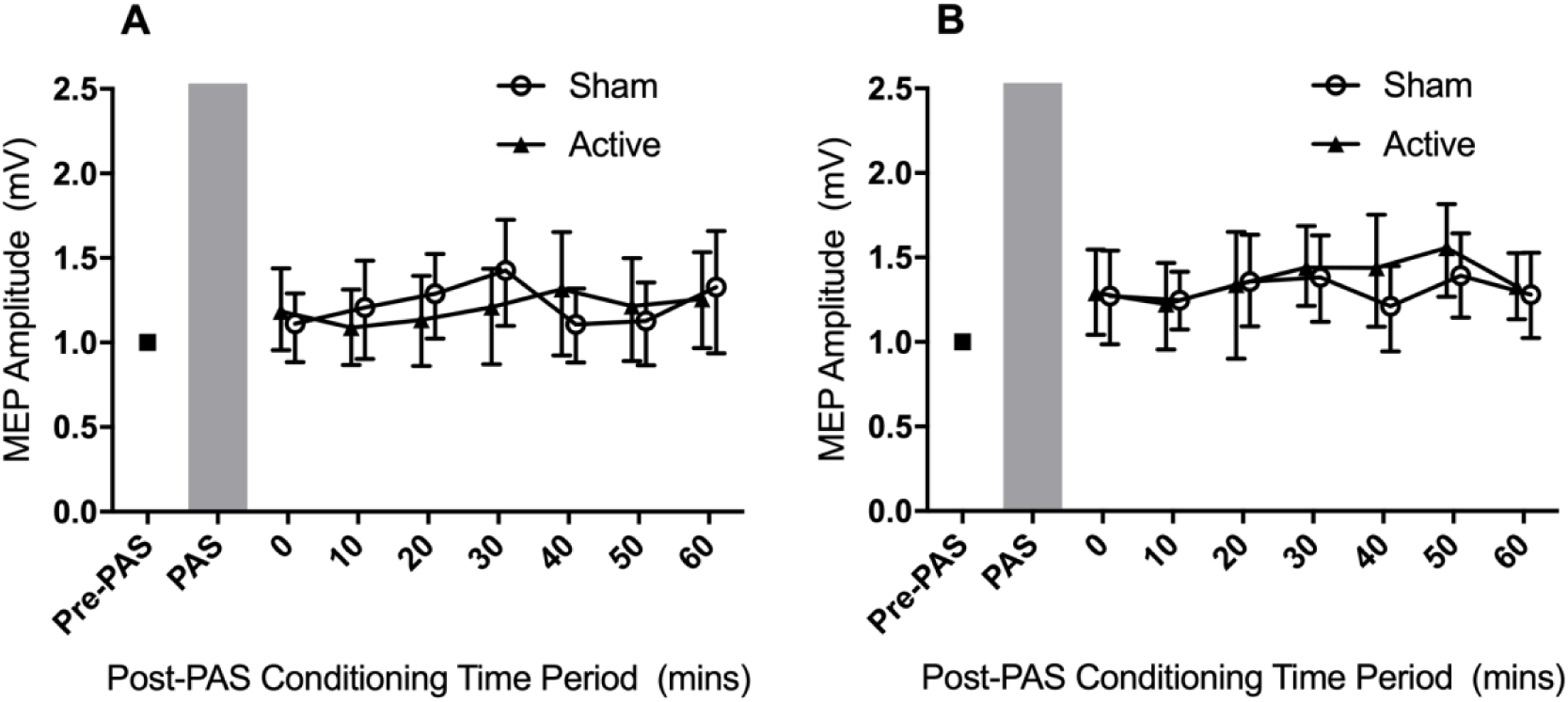
Paired associative stimulation. Group data (estimated marginal means ± bootstrapped 95% confidence intervals) showing the time course of changes in the amplitude of motor evoked potentials (MEPs) immediately after the PAS conditioning stimulus. Post-PAS MEP amplitudes were normalized to pre-PAS amplitudes for each participant. **A)** MEP amplitude changes at baseline, prior to the course of tRNS. **B)** MEP amplitude changes following the 4-week sham-controlled phase.

**Supplementary Figure S6.**
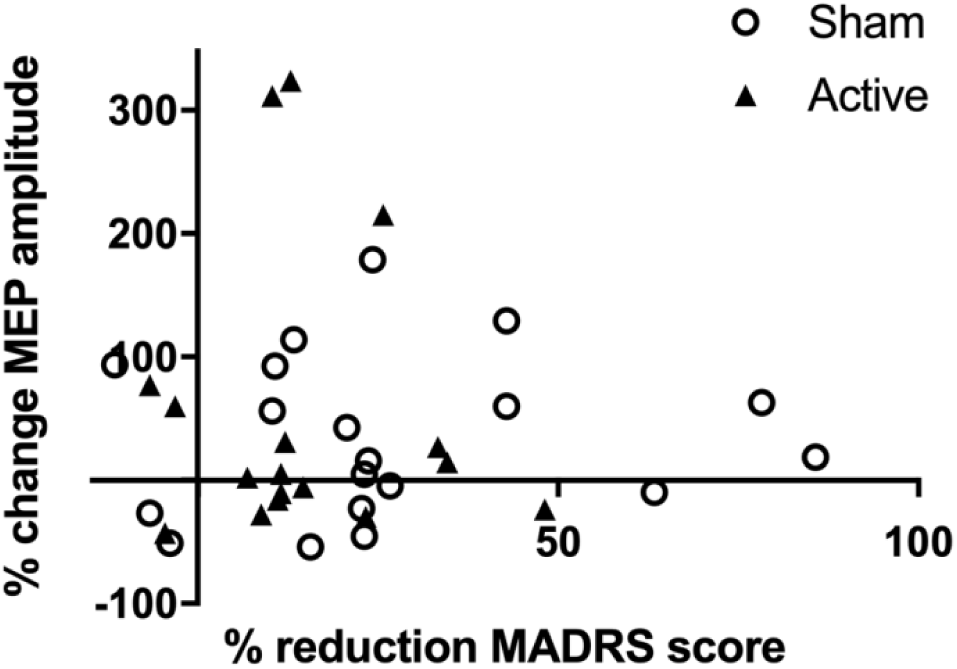
Correlation between mood and neuroplasticity. There was no association between percent change in motor evoked potentials (MEPs) following paired associative stimulation (PAS) and percent change in mood scores over the course of the sham-controlled phase (r = -0.02, *p* = 0.905).

